# Breakfast energy intake and dietary quality and trajectories of cardiometabolic risk factors in older Spanish adults

**DOI:** 10.1101/2024.01.18.24301328

**Authors:** Karla Alejandra Pérez-Vega, Camille Lassale, María Dolores Zomeño, Olga Castañer, Jordi Salas-Salvadó, F. Javier Basterra-Gortari, Dolores Corella, Ramón Estruch, Emilio Ros, Francisco J. Tinahones, Gemma Blanchart, Mireia Malcampo, Daniel Muñoz-Aguayo, Helmut Schröder, Montserrat Fitó, Álvaro Hernáez

## Abstract

**Aims:** To explore the associations between breakfast energy intake and quality and time trajectories of cardiometabolic traits in high cardiovascular risk.

**Methods:** 383 participants aged 55-75 from the PREDIMED-Plus cohort were included. Longitudinal averages of breakfast energy intake and quality were calculated. Three categories were defined for energy intake: 20-30% (reference), <20% (low), and >30% (high). Quality was estimated using the Meal Balance Index; categories were above (reference) or below the median score (low). Smoothed cubic spline mixed effects regressions described trajectories of cardiometabolic indicators (anthropometry, blood pressure, lipids, glucose, glycated hemoglobin, and estimated glomerular filtration rate) at breakfast groups. Inter-group differences in predicted values were estimated by linear regressions.

**Results:** At 36 months, compared to the reference, low- or high-energy breakfasts were associated with differences in: body mass index (low: 0.62 kg/m^2^ [95% confidence interval: 0.28; 0.96]; high: 1.17 kg/m^2^ [0.79; 1.56]), waist circumference (low: 2.24 cm [1.16; 3.32]; high: 4.55 cm [3.32; 5.78]), triglycerides (low: 18.3 mg/dL [15.3; 21.4]; high: 34.5 cm [31.0; 38.1]), and HDL cholesterol (low: −2.13 mg/dL [−3.40; −0.86]; high: −4.56 mg/dL [−6.02; −3.10]). At 36 months, low-quality breakfast was associated with higher waist circumference (1.49 cm [0.67; 2.31]), and triglycerides (3.46 mg/dL [1.13; 5.80]) and less HDL cholesterol (−1.65 mg/dL [−2.61; −0.69]) and glomerular filtration rate (−1.21 mL/min/1.73m^2^ [−2.01; −0.41]).

**Conclusions:** Low- or high-energy and low-quality breakfasts were associated with higher adiposity and circulating triglycerides, and lower HDL cholesterol in high-risk older adults. Low-quality breakfasts were also linked to poorer kidney function.

**LAY SUMMARY:** Our work studied the relationship of the amount of energy consumed at breakfast or the dietary quality of breakfast with the evolution over time of 10 cardiometabolic traits (body mass index, waist circumference, triglycerides, HDL cholesterol, LDL cholesterol, systolic blood pressure, diastolic blood pressure, fasting plasma glucose, glycated hemoglobin, and estimated glomerular filtration rate) in older adults with excess weight and metabolic syndrome.

**Key findings:**

- Compared to a breakfast with an adequate energy intake (containing 20-30% of daily energy), participants consuming either an insufficient or excessive energy in breakfast had higher values of body mass index, waist circumference, and triglycerides, and lower levels of HDL cholesterol.
- Participants with poor breakfast quality, compared to those following a breakfast of higher quality, had higher waist circumference and triglycerides, and lower HDL cholesterol levels and estimated glomerular filtration rate.

**Graphical abstract:** BMI: body mass index; DBP: diastolic blood pressure; eGFR: estimated glomerular filtration rate; Hb1Ac: glycated hemoglobin; HDL-C: high-density lipoprotein cholesterol; LDL-C: low-density lipoprotein cholesterol; SBP: systolic blood pressure; WC: waist circumference.

## INTRODUCTION

Breakfast is a pivotal meal because it breaks the longest fasting time in the day [1]. According to Spanish dietary recommendations, an adequate breakfast provides 20-25% of energy intake [2]. Eating breakfast has been associated with a better quality of the whole diet [3]. Frequent breakfast consumption (three or more times/week, compared to less than three times/week) is related to less risk of obesity, metabolic syndrome, hypertension, type II diabetes, stroke, and cardiovascular mortality [4]. However, only two cross-sectional studies have assessed the relationship between qualitative measurements of breakfast and cardiometabolic health. One study found an association between better breakfast quality and lower values of glycated hemoglobin (Hb1Ac) and better values of a composite cardiometabolic risk score based on high-density lipoprotein cholesterol (HDL-C), low-density lipoprotein cholesterol (LDL-C), triglycerides, and Hb1Ac in older overweight men [5]. The other study, conducted in a general adult population, reported a relationship between high breakfast quality and lower blood pressure, fasting plasma glucose, insulin resistance, total cholesterol, and LDL-C, and risk of being overweight [6]. To the best of our knowledge, no prospective studies have assessed calorie intake in breakfast or the dietary quality of this meal as the exposure. Additionally, none have used repeated measurements of cardiometabolic risk factors over time as the outcome in a well-characterized population.

Our aim was to examine, in older adults with overweight or obesity and metabolic syndrome, the relationship of the amount of energy consumed at breakfast or the dietary quality of breakfast with time-dependent trajectories of a set of cardiometabolic traits: body mass index (BMI), waist circumference (WC), blood triglycerides, HDL-C, LDL-C, systolic blood pressure (SBP), diastolic blood pressure (DBP), fasting plasma glucose, Hb1Ac, and estimated glomerular filtration rate (eGFR).

## METHODS

### Study design and population

This work was performed in a subsample of individuals recruited in the Prevención con Dieta Mediterránea-Plus (PREDIMED-Plus) study, and we used the study data for a set of observational analyses. PREDIMED-Plus is a randomized clinical trial that compares the effect of a lifestyle intervention with an energy reduced Mediterranean diet (MedDiet) plus physical activity with an *ad libitum* MedDiet without advice on exercise (control group) on the incidence of cardiovascular disease [7]. Eligible participants were women between 60–75 years and men between 55–75 years with BMI between 27 and 40 kg/m^2^ and at least three criteria for metabolic syndrome: 1) triglycerides ≥150 mg/dL or triglyceride-lowering medication; 2) fasting glucose ≥100 mg/dL or glucose-lowering medication; 3) SBP/DBP ≥130/85 mmHg or antihypertensive medication; 4) HDL-C <50 mg/dL in women and <40 mg/dL in men; and/or 5) WC ≥88 cm in women and ≥102 cm in men [7]. Complementary information of the protocol (setting, locations, relevant dates, periods of recruitment, follow-up) and details of the intervention are available elsewhere [7,8]. Participants in the two arms of the study received no instructions on how to prepare breakfast other than structuring it following a MedDiet. They were advised to consume low-fat dairy products, whole grain cereal or bread, a protein rich food, extra virgin olive oil and/or nuts as a source of fat, and a fresh seasonal fruit, and to avoid ultra-processed foods [7,9]. Participants in both arms of the trial experienced weight loss in the first 12 months of the study and an associated improvement in some parameters such as lipid profile and blood pressure, although the improvements were significantly greater in the energy reduced MedDiet group [8].

This sub-study was conducted in PREDIMED-Plus participants recruited at Hospital del Mar Research Institute (Barcelona, Spain) who had completed at least one three-day food record (**Figure 1**). Our analyses are reported following the guidelines described by the STrengthening the Reporting of OBservational studies in Epidemiology statement.

**Figure 1.**
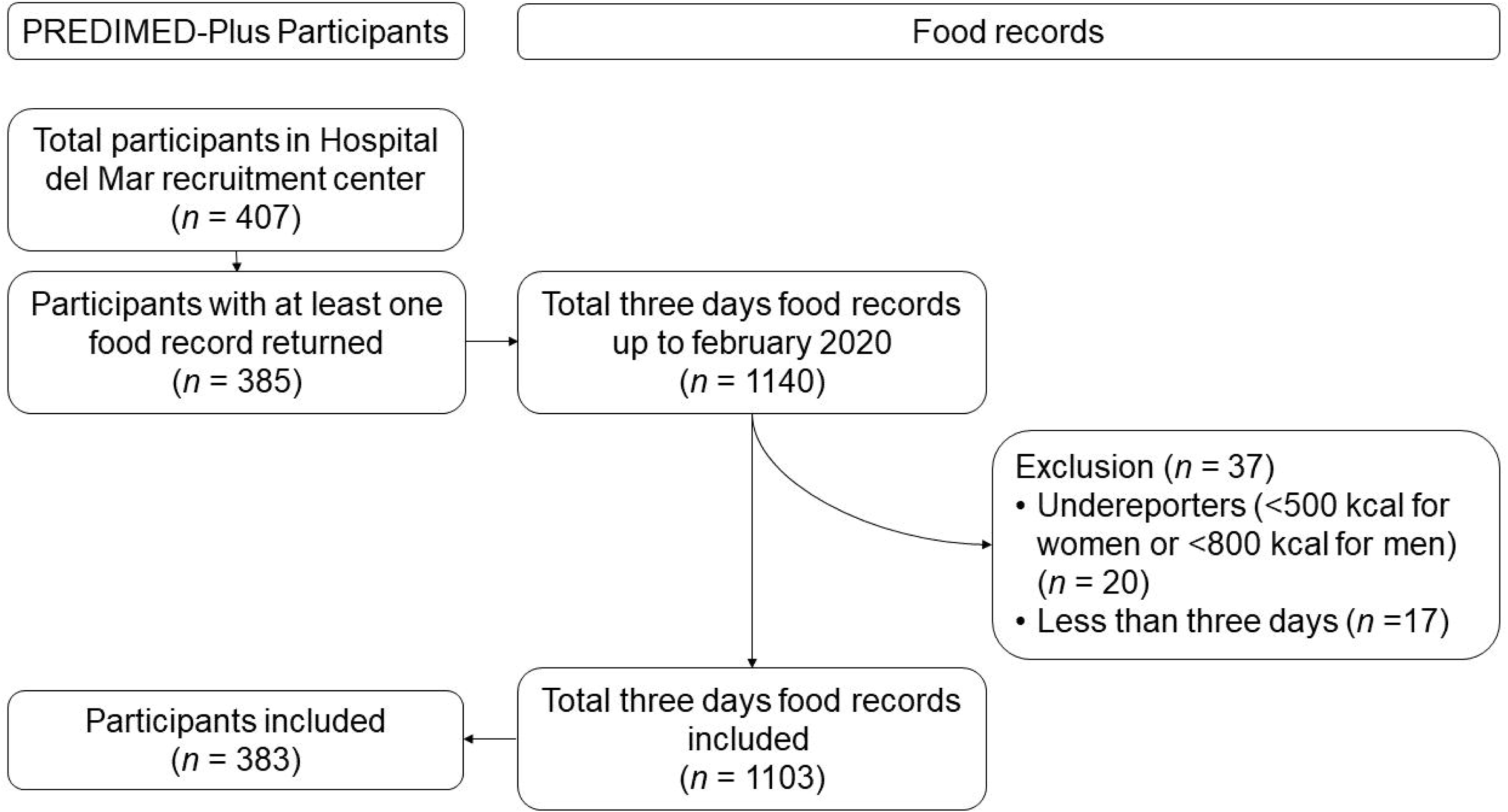
Study flow chart

### Breakfast data

We first assessed dietary intake with three-day food records at three time points: baseline, 24 months, and 36 months of follow-up. Before each visit, a nutritionist facilitated a pre-structured form to record everything the participant ate and drank in the following meals: breakfast, morning snack, lunch, afternoon snack, dinner, and night snack. Participants were instructed to self-report consumption of all foods and beverages in two labor days and one weekend day, with detailed descriptions using household measures or weighted food, and explain the ingredients in recipes or brands of processed food. Trained nutritionists reviewed the food records together with the participants to check for completeness, searching particularly unrecorded items such as sugar, bread, oil, or butter. Reviewed food records were computerized and analyzed in the PCN Pro 1.0 software [10] (University of Barcelona, Barcelona, Spain), with Spanish-specific nutritional composition table [11]. We obtained total energy (kcal) and macronutrients (g) from the whole day and separately for breakfast. We considered as breakfast any food or beverage reported before lunch and registered as breakfast and/or morning snack. We included the morning snack as part of the breakfast because a relevant proportion of our population divided it into two meals, one of them being lighter but still considered a breakfast (for example, coffee with milk upon waking up and a mid-morning sandwich). Food records with an average daily energy intake of <500 and >3,500 kcal for women or <800 and >4,200 for men [12] were discarded and participants with less than one full record (three days) were excluded. For sensitivity analyses, we also estimated the proportion of energy intake at breakfast and mid-morning snack.

We used these data to calculate the proportion of energy consumed at breakfast relative to the total daily energy intake. We also used it to estimate the breakfast quality using the Meal Balance Index [13]. This score informs of the quality of a meal according to the content of nine nutrients (proteins, total fat, fiber, potassium, calcium, iron, sodium, added sugars and saturated fat). It uses: 1) Acceptable Macronutrient Distribution Ranges as reference for proteins and fats; 2) Daily Values for fiber, potassium, calcium, and iron; and 3) World Health Organization recommendations for proportions of added sugars, saturated fats, and sodium. We estimated the amount of the nutrients ingested at breakfast and expressed it per 2,000 kcal, compared it to the reference values, and assigned a score (ranging from 0 to 100 for each nutrient) according to these levels. The translation of the intake values of the nine nutrients or food groups into scores is described in **Table 1**. Finally, we calculated the breakfast quality score as the weighted average of the nine nutrient/food group scores (scores for potassium and saturated fat weighed double). Total score ranged from 0 to 100. Higher scores mean greater quality of the meal [13].

**Table 1.** Intake values for the calculation of the Meal Balance Score.

|  | Score 0 | 0 to 100 points<br>(increasing,<br>proportional<br>values) | Score 100 | 100 to 0 points<br>(decreasing,<br>proportional<br>values) | Score 0 |
| --- | --- | --- | --- | --- | --- |
| Protein (%) | ≤5 | >5 to <10 | ≥10 to ≤35 | >35 to <52.5 | ≥52.5 |
| Total fat (%) | ≤10 | >10 to <20 | ≥20 to ≤35 | >35 to <52.5 | ≥52.5 |
| Fiber<br>(g/2,000 kcal) | ≤14 | >14 to <28 | ≥28 to ≤56 | >56 to <84 | ≥84 |
| Potassium<br>(mg/2,000 kcal) | ≤1750 | >1,750<br>to <3,500 | ≥3,500<br>to ≤7,000 | >7,000<br>to <10,500 | ≥10,500 |
| Calcium<br>(mg/2,000kcal) | ≤500 | >500<br>to <1,000 | ≥1,000<br>to ≤2,000 | >2,000<br>to <3,000 | ≥3,000 |
| Iron<br>(mg/2,000 kcal) | ≤9 | >9 to <18 | ≥18 to ≤36 | >36 to <54 | ≥54 |
| Added sugar (%) | - | - | 0 to ≤10 | >10 to <15 | ≥15 |
| Saturated fat (%) | - | - | 0 to ≤10 | >10 to <15 | ≥15 |
| Sodium<br>(mg/2,000kcal) | - | - | 0<br>to ≤2,000 | >2,000<br>to <3,000 | ≥3,000 |

Adapted from Mainardi F et al., Plos One, 2020 (DOI: 10.1371/journal.pone.0244391)

### Cardiometabolic risk factors

Healthcare professionals measured weight, height and WC using calibrated equipment and following the study protocol (www.predimedplus.com) [8]. Participants’ weight was recorded without shoes and with light clothing using a calibrated high-quality electronic scale. Height was measured with a calibrated stadiometer at the beginning of the study. BMI was calculated as weight (kg) divided by height squared (m^2^). WC was determined in the midpoint between the lowest rib and the iliac crest using an anthropometric tape. Blood pressure (BP) was measured in triplicate using a calibrated automated oscillometer (Omron HEM-705CP, Netherlands) with participants seated and after five minutes of rest, and the mean of the three measurements was calculated [8].

We collected fasting ethylenediaminetetraacetic acid plasma at baseline and in the follow-up visits at six, 12, and 36 months and measured triglycerides (Triglycerides CP, Horiba ABX), total cholesterol (Cholesterol CP, Horiba ABX), HDL-C (HDL Direct CP, Horiba ABX), glucose (Glucose HK CP, Horiba ABX), HbA1c (HbA1c WB, Horiba ABX), and creatinine (Creatinine 120 CP, Horiba ABX) in an autoanalyzer ABX Pentra (Horiba ABX SAS, Spain). We calculated LDL-C with the Friedewald formula only when triglycerides were <300 mg/dL, higher values (≥300 mg) implied a missing value for LDL-C. eGFR was estimated using plasma creatinine, sex, and age in the equation for European population [14].

### Other variables

Healthcare professionals collected data at baseline on: age, sex, educational level (elementary school, middle/high school or higher education), smoking habit (never smoker, current smoker or former smoker), and prevalence of diabetes, hypercholesterolemia, and hypertension as previously described [8].

### Ethical aspects

This study follows the Declaration of Helsinki for Medical Research on human subjects. Before the study started, local institutional ethic committees approved the protocol. All participants signed an informed consent before enrolling in the study. The protocol was registered in the ISRCTN Registry (PREDIMED-Plus: ISRCTN89898870). We followed the EQUATOR Network principles for guidance on study ethics and reporting.

### Statistical analysis

We described normally distributed continuous variables using means and standard deviations (SD), non-normally distributed continuous variables using medians and 1^st^-3^rd^ quartiles, and categorical variables as proportions. We analyzed the association between the percentage of energy consumed at breakfast and the breakfast quality score by a Spearman’s Rank correlation coefficient.

We evaluated the association of energy intake at breakfast or the dietary quality of breakfast with time-dependent trajectories of cardiometabolic risk factors. We first calculated the longitudinal average of the percentage of energy consumed at breakfast and the breakfast quality score through all food records available for a given participant. We then defined three categories according to the longitudinal average of the breakfast energy intake: 20-30% (reference group), <20% (low intake), and >30% (high intake). Although recommendations suggest 20-25% of daily energy intake for breakfast, we widened the range up to 30% to consider morning snacks. Similarly, we defined two categories according to breakfast quality: score above the median (reference group) and below the median (low quality). We modeled the trajectories of each cardiometabolic risk factor using smoothed cubic spline mixed effects regression models, including an interaction term between age at every follow-up visit (as the time variable) and breakfast-related groups to allow for different trajectories in participants in the different groups [15]. Analyses were adjusted for age, sex, PREDIMED-Plus intervention group, educational level, smoking, and total daily intake of kilocalories. Analyses that used lipid profile biomarkers as outcomes were further adjusted for prevalence of hypercholesterolemia at baseline, those that assessed BP were adjusted for hypertension at baseline, and those on glucose and Hb1Ac were adjusted for diabetes at baseline. Analyses on breakfast energy intake groups were further adjusted for breakfast quality, and those on breakfast quality groups were further adjusted for the percentage of energy consumed at breakfast. We used predicted values to plot mean trajectories in the different groups. We calculated the mean inter-group differences in cardiometabolic risk factors at baseline, six, 12, 18, 24, 30 and 36 months using linear regressions.

Analyses were performed in R Software, version 4.1.2. The code for these analyses is available in: https://github.com/alvarohernaez/Breakfast_trajectories/.

## RESULTS

### Study population

Our study subjects were 383 participants of the PREDIMED-Plus study with available and plausible 1,103 diet records (**Figure 1**). By study design, all participants were older adults (51.4% women), had overweight (19.3%) or obesity (80.7%), and harbored the metabolic syndrome. Consequently, participants presented a high prevalence of cardiovascular risk factors (**Table 2**). We found no clinically meaningful differences in baseline characteristics among participants in different breakfast energy intake groups and different breakfast quality categories.

**Table 2.**
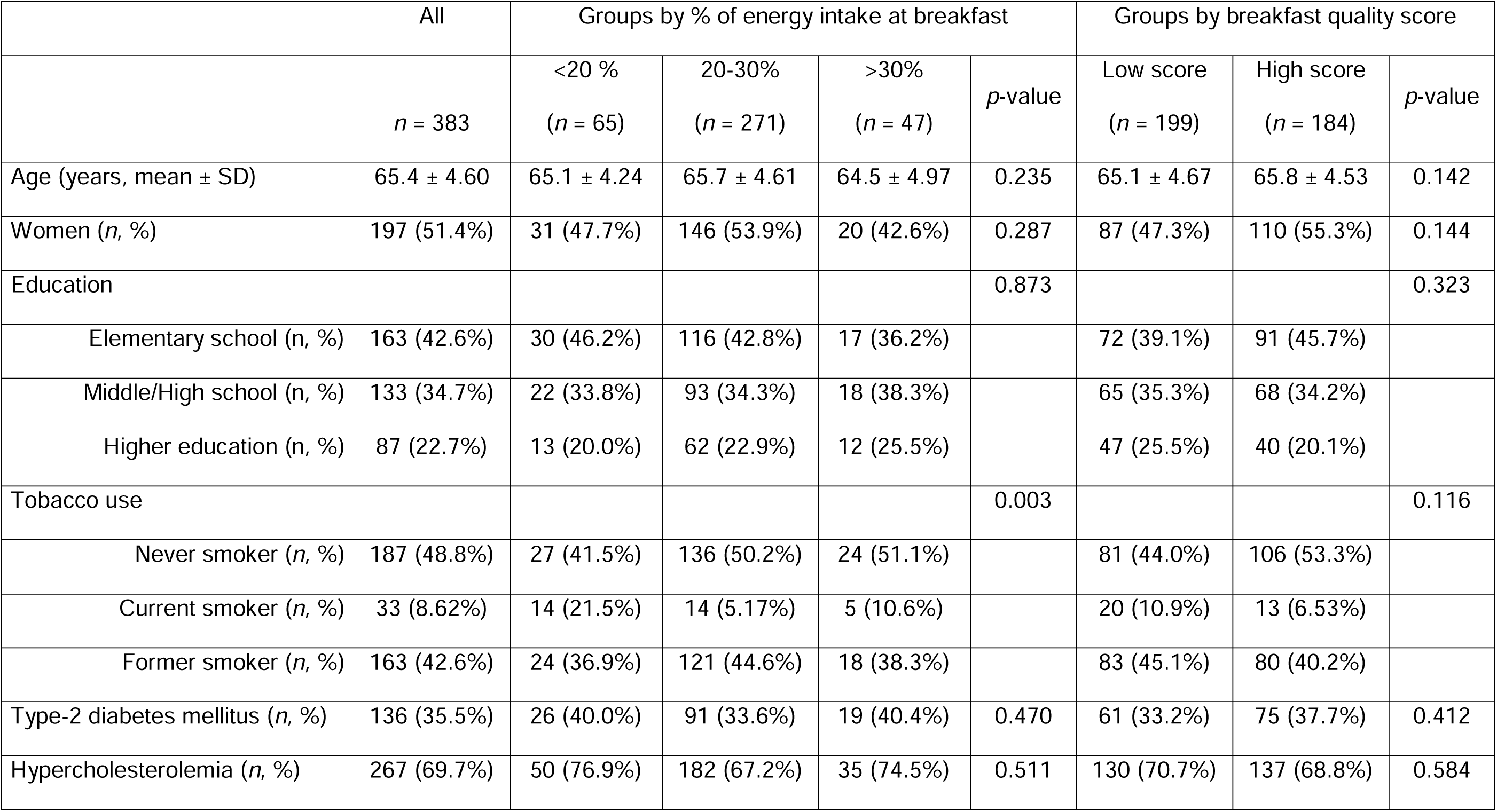
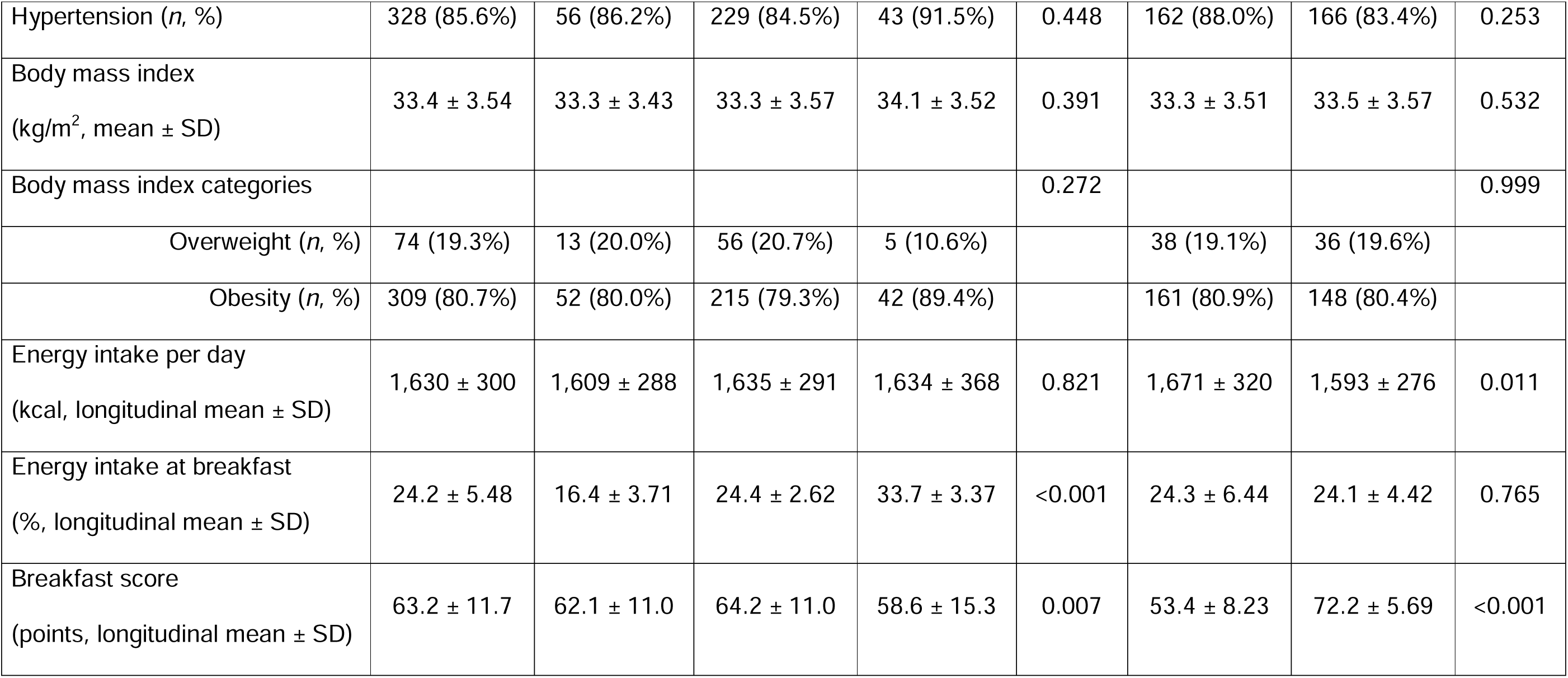
Characteristics of study participants.

|  | All | Groups by % of energy intake at breakfast |  |  |  | Groups by breakfast quality score |  |  |
| --- | --- | --- | --- | --- | --- | --- | --- | --- |
|  | *n* = 383 | <20 %<br>(*n* = 65) | 20-30%<br>(*n* = 271) | >30%<br>(*n* = 47) | *p*-value | Low score<br>(*n* = 199) | High score<br>(*n* = 184) | *p*-value |
| Age (years, mean ± SD) | 65.4 ± 4.60 | 65.1 ± 4.24 | 65.7 ± 4.61 | 64.5 ± 4.97 | 0.235 | 65.1 ± 4.67 | 65.8 ± 4.53 | 0.142 |
| Women (*n*, %) | 197 (51.4%) | 31 (47.7%) | 146 (53.9%) | 20 (42.6%) | 0.287 | 87 (47.3%) | 110 (55.3%) | 0.144 |
| Education |  |  |  |  | 0.873 |  |  | 0.323 |
| Elementary school (*n*, %) | 163 (42.6%) | 30 (46.2%) | 116 (42.8%) | 17 (36.2%) |  | 72 (39.1%) | 91 (45.7%) |  |
| Middle/High school (*n*, %) | 133 (34.7%) | 22 (33.8%) | 93 (34.3%) | 18 (38.3%) |  | 65 (35.3%) | 68 (34.2%) |  |
| Higher education (*n*, %) | 87 (22.7%) | 13 (20.0%) | 62 (22.9%) | 12 (25.5%) |  | 47 (25.5%) | 40 (20.1%) |  |
| Tobacco use |  |  |  |  | 0.003 |  |  | 0.116 |
| Never smoker (*n*, %) | 187 (48.8%) | 27 (41.5%) | 136 (50.2%) | 24 (51.1%) |  | 81 (44.0%) | 106 (53.3%) |  |
| Current smoker (*n*, %) | 33 (8.62%) | 14 (21.5%) | 14 (5.17%) | 5 (10.6%) |  | 20 (10.9%) | 13 (6.53%) |  |
| Former smoker (*n*, %) | 163 (42.6%) | 24 (36.9%) | 121 (44.6%) | 18 (38.3%) |  | 83 (45.1%) | 80 (40.2%) |  |
| Type-2 diabetes mellitus (*n*, %) | 136 (35.5%) | 26 (40.0%) | 91 (33.6%) | 19 (40.4%) | 0.470 | 61 (33.2%) | 75 (37.7%) | 0.412 |
| Hypercholesterolemia (*n*, %) | 267 (69.7%) | 50 (76.9%) | 182 (67.2%) | 35 (74.5%) | 0.511 | 130 (70.7%) | 137 (68.8%) | 0.584 |
| Hypertension (n, %) | 328 (85.6%) | 56 (86.2%) | 229 (84.5%) | 43 (91.5%) | 0.448 | 162 (88.0%) | 166 (83.4%) | 0.253 |
| Body mass index<br>(kg/m<sup>2</sup>, mean ± SD) | 33.4 ± 3.54 | 33.3 ± 3.43 | 33.3 ± 3.57 | 34.1 ± 3.52 | 0.391 | 33.3 ± 3.51 | 33.5 ± 3.57 | 0.532 |
| Body mass index categories |  |  |  |  | 0.272 |  |  | 0.999 |
| Overweight (n, %) | 74 (19.3%) | 13 (20.0%) | 56 (20.7%) | 5 (10.6%) |  | 38 (19.1%) | 36 (19.6%) |  |
| Obesity (n, %) | 309 (80.7%) | 52 (80.0%) | 215 (79.3%) | 42 (89.4%) |  | 161 (80.9%) | 148 (80.4%) |  |
| Energy intake per day<br>(kcal, longitudinal mean ± SD) | 1,630 ± 300 | 1,609 ± 288 | 1,635 ± 291 | 1,634 ± 368 | 0.821 | 1,671 ± 320 | 1,593 ± 276 | 0.011 |
| Energy intake at breakfast<br>(%, longitudinal mean ± SD) | 24.2 ± 5.48 | 16.4 ± 3.71 | 24.4 ± 2.62 | 33.7 ± 3.37 | <0.001 | 24.3 ± 6.44 | 24.1 ± 4.42 | 0.765 |
| Breakfast score<br>(points, longitudinal mean ± SD) | 63.2 ± 11.7 | 62.1 ± 11.0 | 64.2 ± 11.0 | 58.6 ± 15.3 | 0.007 | 53.4 ± 8.23 | 72.2 ± 5.69 | <0.001 |

Average energy intake at breakfast was 23% at baseline, 24% at 24 months of follow-up, and 25% at 36 months of follow-up. We found no association between the percentage of energy consumed at breakfast and breakfast quality (*r* = −0.037, *p*-value = 0.47).

### Breakfast and adiposity

Participants with low and high breakfast energy intake showed increasing values of BMI over time compared to the reference group (inter-group difference at 36 months, low energy intake: +0.62 kg/m^2^, 95% confidence interval (CI) 0.28 to 0.96; high energy intake: +1.17 kg/m^2^, 95% CI 0.79 to 1.56; **Figures 2A-2C**). No sustained inter-group differences according to breakfast quality were found (**Figures 2D-2E**).

**Figure 2.**
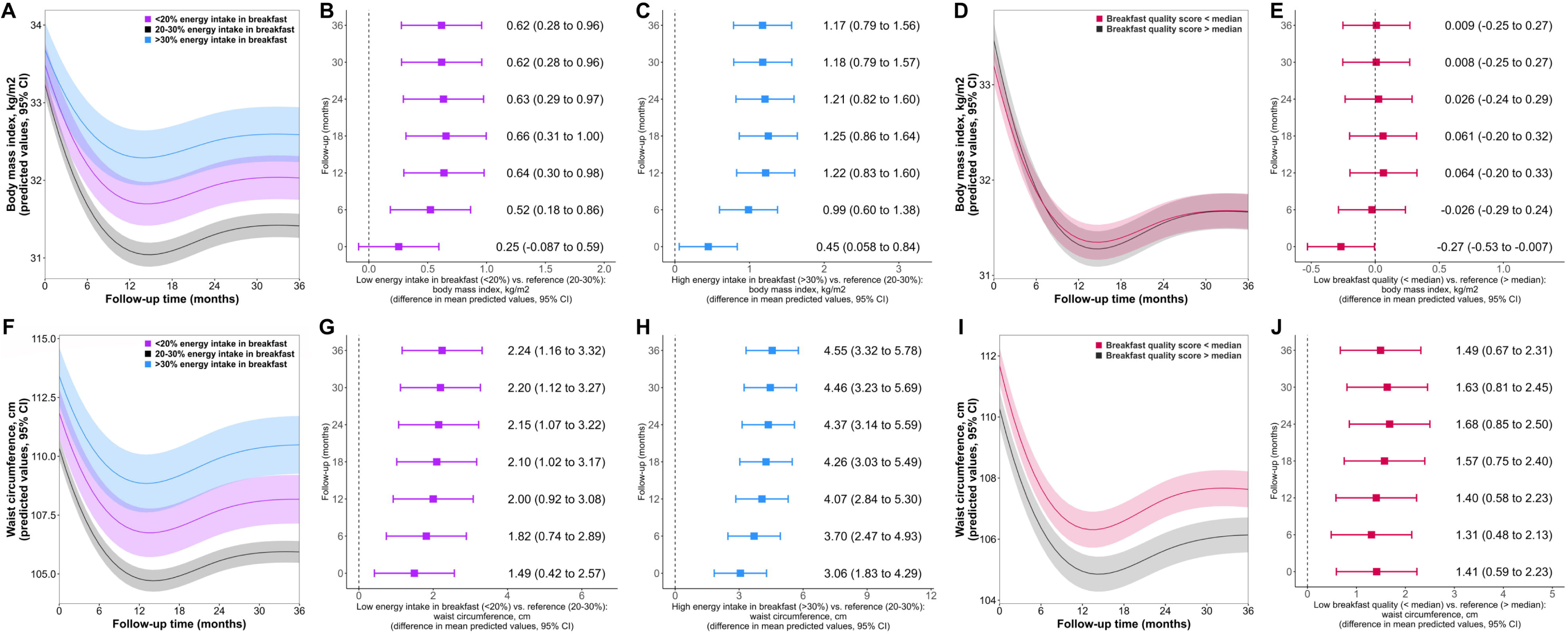
Trajectories at breakfast energy intake groups and breakfast quality groups plus inter-group differences, for body mass index (A-E) and waist circumference (F-J).

**Figure 3.**
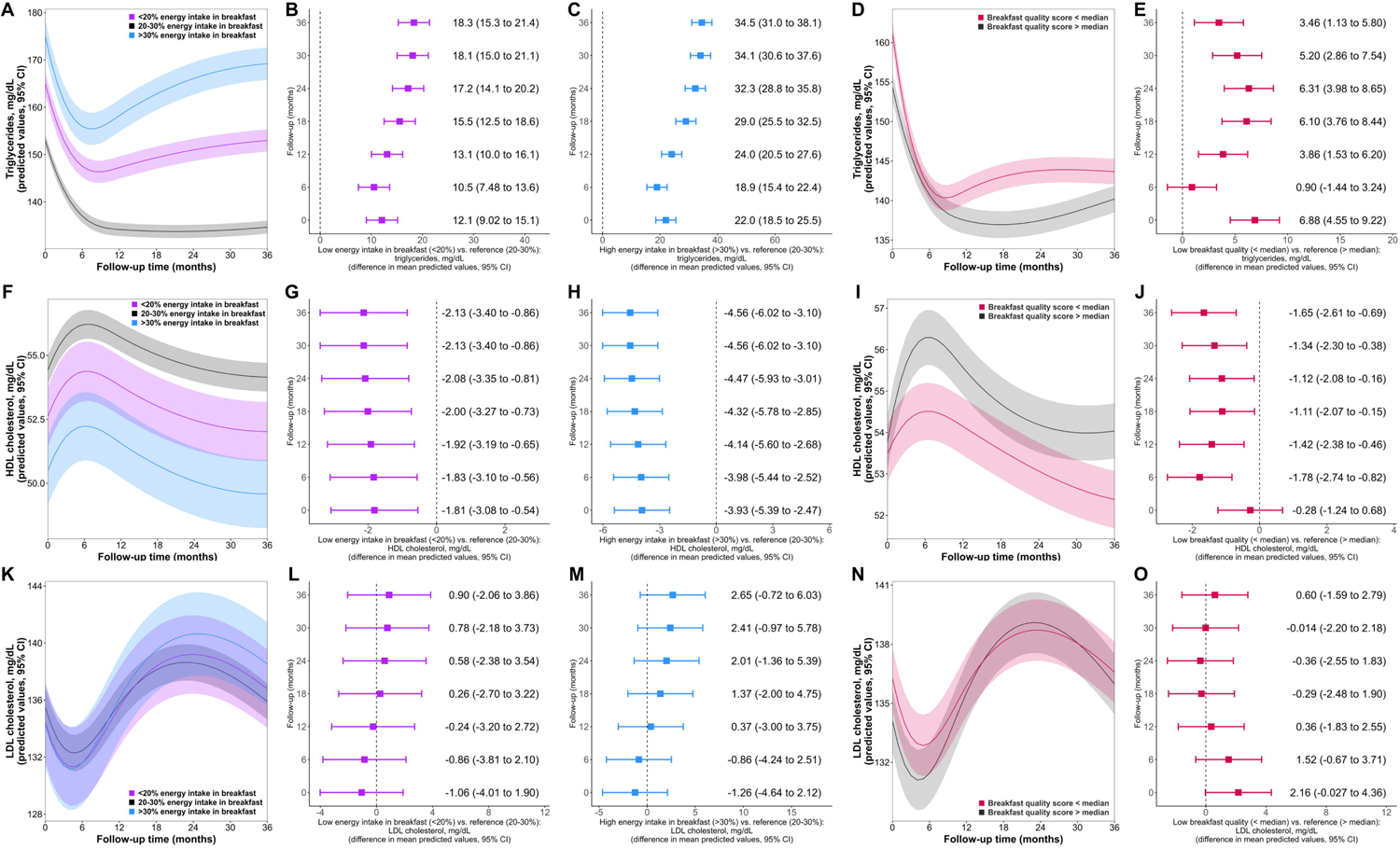
Trajectories at breakfast energy intake groups and breakfast quality groups plus inter-group differences, for triglycerides (A-E), high-density lipoprotein cholesterol (F-J) and for low-density lipoprotein cholesterol (K,O).

Participants with low and high energy intake at breakfast showed a more pronounced rebound in WC values after the first year of the study and increasing differences over time (inter-group difference at 36 months: low energy intake: +2.24 cm, 95% CI 1.16 to 3.32; high energy intake: +4.55 cm, 95% CI 3.32 to 5.78; **Figures 2F-2H**). Participants with low breakfast quality also showed higher WC (inter-group differences at 36 months: +1.49 cm, 95% CI 0.67 to 2.31) (**Figures 2I-2J**).

### Breakfast and lipid profile

Triglyceride trajectories were different depending on breakfast groups. Participants with low and high energy intake at breakfast showed a rebound in triglyceride values after six months of follow-up (particularly for those with high energy intake) that was not evident under the reference energy consumption at breakfast (**Figure 3A**). Triglyceride values were higher and inter-group differences grew over time (inter-group difference at 36 months, low energy intake: +18.3 mg/dL, 95% CI 15.3 to 21.4; high energy intake: +34.5 mg/dL, 95% CI 31.0 to 38.1; **Figures 3B-3C**). Participants with low breakfast quality also showed an early rebound in triglyceride concentrations after the decrease in the first months of the PREDIMED-Plus intervention (**Figure 3D**) and higher mean triglyceride values (inter-group difference at 36 months: +3.46 mg/dL, 95% CI 1.13 to 5.80) (**Figure 3E**).

The shape of HDL-C trajectories in all breakfast groups was similar, but predicted mean HDL-C levels were consistently lower in both low and high breakfast energy groups compared to the reference group (inter-group difference at 36 months, low energy intake: −2.13 mg/dL, 95% CI −3.40 to −0.86; high energy intake: −4.56 mg/dL, 95% CI −6.02 to −3.10; **Figures 3F-3H**). Predicted mean HDL-C concentrations were also lower in participants with low breakfast quality (inter-group difference at 36 months, −1.65 mg/dL, 95% CI −2.61 to −0.69) (**Figures 3I-3J**).

LDL-C trajectories were comparable across breakfast energy intake groups and breakfast quality groups, and no inter-group differences were observed (**Figures 3K-3O**).

### Breakfast and blood pressure

There were no differences in the SBP trajectories according to energy intake at breakfast (**Figures 4A-4C**). Regarding breakfast quality, slightly higher mean predicted values of SBP were observed at 12-18 months of follow-up in participants with low breakfast quality (**Figures 4D-4E**). Similarly, DBP trajectories were comparable for energy intake groups (**Figures 4F-4H**) and slightly higher mean predicted values of DBP were reported at 12-18 months in participants with low breakfast quality (**Figures 4I-4J**).

**Figure 4.**
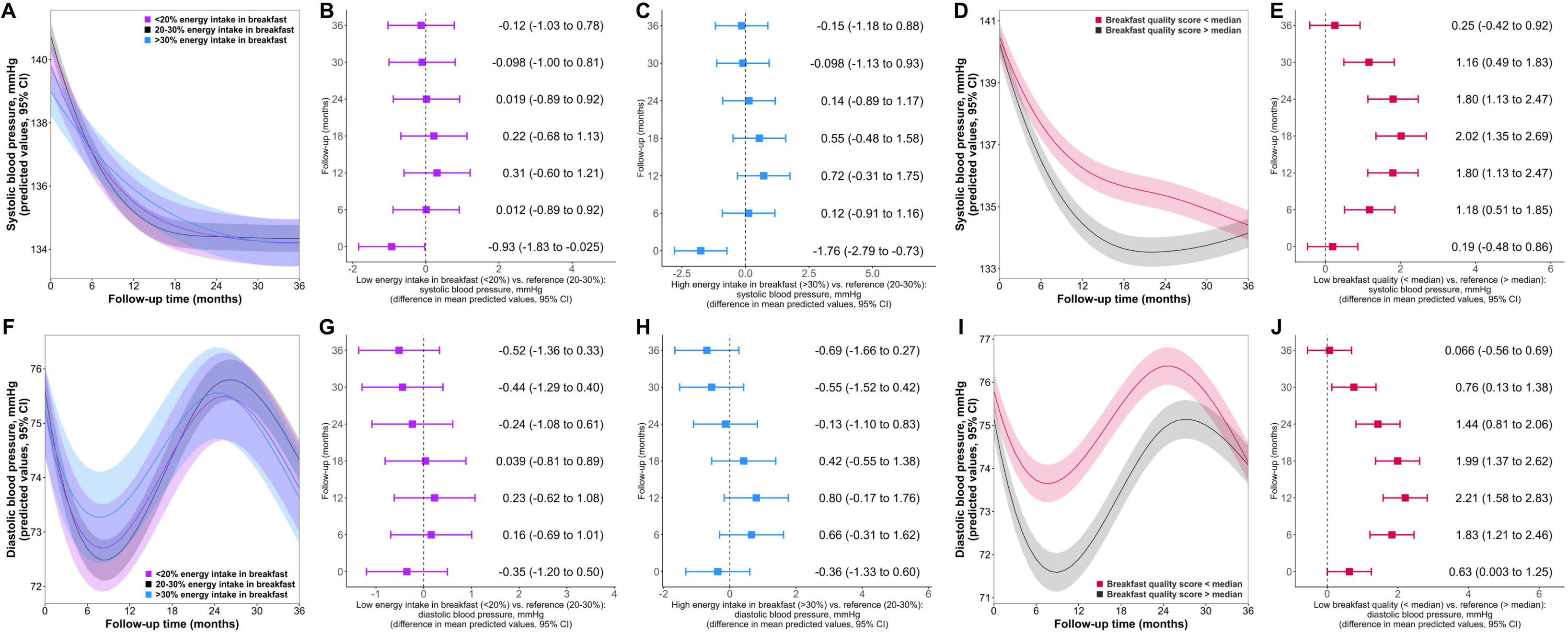
Trajectories at breakfast energy intake groups and breakfast quality groups plus inter-group differences, for systolic blood pressure (A-E) and diastolic blood pressure (F-J).

### Breakfast and glucose metabolism

Participants in the reference group for energy intake at breakfast disclosed a more pronounced decrease in fasting plasma glucose values during the first 6 months of the study compared to participants with low and high energy intakes at breakfast. However, glucose levels were not different between groups at ensuing follow-up points (**Figures 5A-5C**). Trajectories in groups according to breakfast quality were comparable (**Figures 5D-5E**). Hb1Ac trajectory curves were similar among groups of breakfast energy intake and breakfast quality (**Figures 5F-5J**). Nonetheless, fasting plasma glucose and Hb1Ac values were slightly higher in participants with a low-quality breakfast, although differences were neither significant nor clinically relevant.

**Figure 5.**
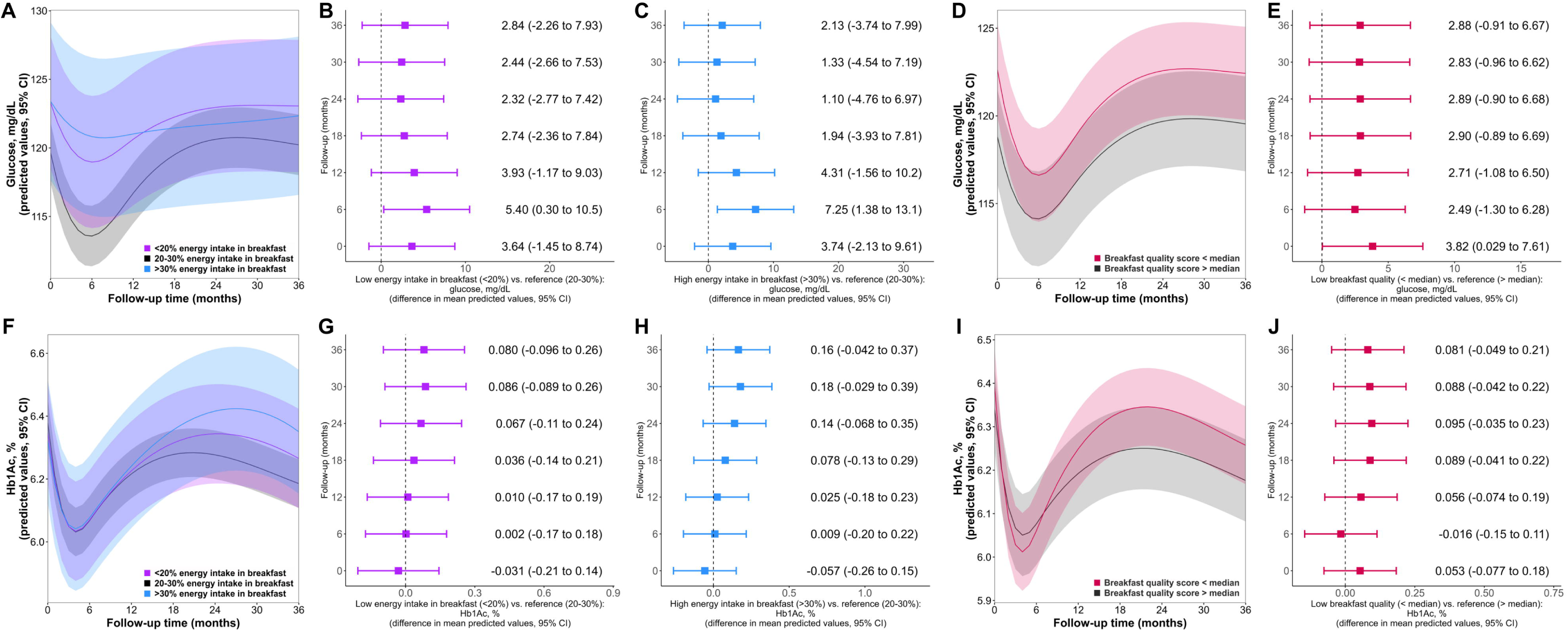
Trajectories at breakfast energy intake groups and breakfast quality groups plus inter-group differences, for fasting plasma glucose (A-E) and glycated hemoglobin (F-J).

### Breakfast and estimated glomerular filtration rate

eGFR trajectories in the groups of energy intake had a similar shape (**Figures 6A-6C**). In relation to breakfast quality, participants in the group with a low-quality breakfast had lower mean predicted eGFR (inter-group differences at 36 months, −1.21 mL/min/1.73m^2^, 95% CI −2.01 to −0.41; **Figures 6D-6E**).

**Figure 6.**
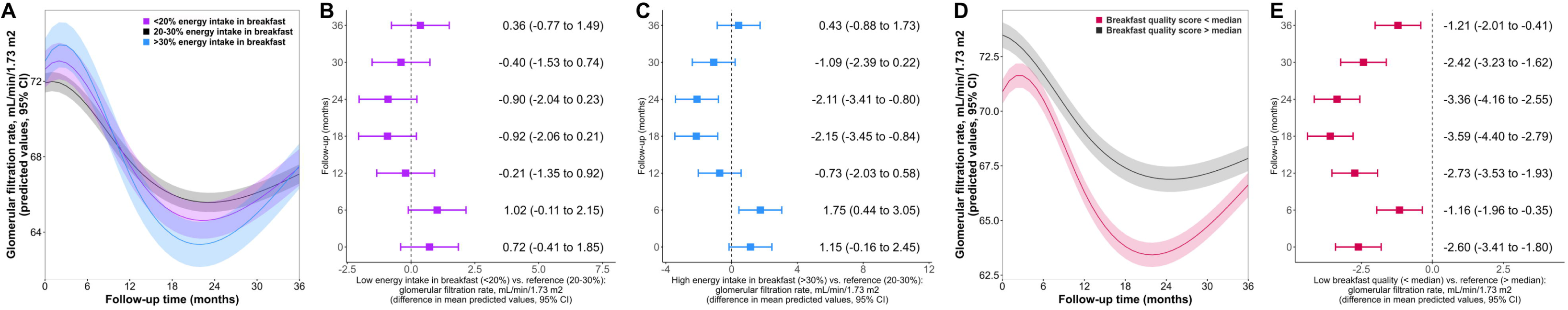
Trajectories at breakfast energy intake groups and breakfast quality groups plus inter-group differences, for glomerular filtration rate.

## DISCUSSION

In older adults at high cardiovascular risk, the energy consumed at breakfast and its nutritional quality are linked to differences in cardiometabolic health. Compared to a breakfast containing 20-30% of daily energy intake, participants consuming either low or high energy breakfasts displayed higher values of BMI, WC, and circulating triglycerides, and lower HDL-C. Additionally, they showed a rebound in WC and triglycerides after the first year of intervention that was not evident in participants with an adequate energy intake at breakfast. When focusing on the quality of breakfast, participants with poor breakfast quality also had higher WC and triglycerides and lower HDL-C levels and eGFR than those with a higher breakfast quality.

Our findings show that an insufficient energy intake at breakfast is associated with greater adiposity, which concurs with previous evidence. Adults consuming less than 22% of their daily energy at breakfast in a cohort study had a higher BMI regardless of their total intake of energy when compared to consumers of higher intakes [16]. In a retrospective cross-sectional study, men who ate a small breakfast had higher BMI than those who had standard or large breakfasts [17]. Finally, in a clinical trial involving women participating in a 12-week isocaloric weight loss program, those who consumed 14% of energy intake at breakfast and 50% at dinner achieved less weight loss and lower decreases in WC compared to those who had 50% at breakfast and 14% at dinner [18]. Eating breakfast has been linked to increased satiety, which in turn leads to reduced total energy intake [19] and greater postprandial thermogenesis [20], providing a possible mechanism for less adiposity. On the other hand, our results on an association between high energy intakes at breakfast (>30%) with greater adiposity are novel. Unlike previous studies, we distinguished between reference and high energy intakes at breakfast and adjusted our analyses for the total energy intake in the day and the quality of the breakfast, which may explain our capacity to detect these differences. Having 20-30% of daily calories for breakfast was also associated with favorable changes in other cardiovascular risk factors related to adiposity, such as lower levels of triglycerides (triglyceride differences were clinically relevant, up to 36 mg/dL) and higher concentrations of HDL-C. These results align with one cross-sectional study reporting that participants eating breakfast had lower levels of triglycerides and higher HDL-C than those skipping breakfast [21].

In terms of the quality of breakfast, higher scores were also associated with lower adiposity (lower WC). While the ideal breakfast composition is debatable, our findings are in line with another study suggesting that people who choose to consume fruit, unprocessed and unsweetened cereal flakes, nuts, and yogurt for breakfast tend to have lower abdominal obesity [22]. Breakfast quality could modulate factors that may impact adiposity, as a high-quality breakfast (rich in protein and carbohydrates) decreased appetite, cravings, and postprandial ghrelin levels in a randomized controlled trial with obese adults [23]. Our study is the first to associate a high-quality breakfast with lower triglyceride and higher HDL-C levels, something that can be explained by the association between lower adiposity and a better triglyceride and HDL-C status [24]. Besides, we also observed for the first time, that participants following a high-quality breakfast had higher eGFR than people in the low-quality breakfast group. Evidence on breakfast and renal function is mainly focused on studies about skipping breakfast, as adults who omitted breakfast had greater odds of chronic kidney disease and proteinuria in cross-sectional studies [25,26]. Lower adiposity in individuals with a high-quality breakfast may explain better kidney function [27].

We observed no clear differences in BP according to energy intake at breakfast, despite the slightly higher BP levels in participants with low-quality breakfast in some time points. These differences were not clinically relevant (≤3 mmHg), as opposed to those observed for BMI, WC, triglycerides and HDL-C. Compared to skipping breakfast, eating breakfast has been associated with lower SBP (differences of ≤5 mmHg) and DBP (differences of ≤2 mmHg) in previous studies [28,29]. We also observed no clear differences for fasting plasma glucose or Hb1Ac levels, apart from a more pronounced decrease in glucose values during the first six months of the study in participants who consumed 20-30% of daily calories at breakfast and a non-clinically relevant difference in fasting plasma glucose and Hb1Ac values in those with a low-quality breakfast. These slight differences could be explained by the greater content of fiber in a healthy breakfast, which could delay the absorption of carbohydrates and optimize insulin sensitivity through a wide range of molecular mechanisms [30]. The lack of robust differences in parameters related to glucose metabolism does not concur with previous studies that have reported an increased risk of developing type II diabetes among adults who skip breakfast [31–33]. Discrepancies between previous findings and our results can be explained by the different definition of exposure (previous studies are focused on skipping breakfast and our exposures were energy consumed at breakfast or breakfast quality), the fact that some of these studies were cross-sectional, and that their participants were younger and had fewer cardiovascular risk factors [31–33].

Our study had some limitations. First, this study is observational, and we do not know whether the associations between the quantity and quality of breakfast and the risk factors trajectories of breakfast are causal or whether they may be explained by residual confounding. We tried to minimize this source of bias by adjusting for several covariates (e.g., age, sex, intervention group, education level, smoking habit, total daily intake of energy, and diet quality). Nevertheless, these relationships should be verified in future nutritional intervention studies. Second, nutritional assessment was based on three-day food diaries. Although it is the gold standard, it may imply some bias due to the subjective nature of participants’ self-reporting. We tried to minimize this limitation by reviewing the food records with the participants and by excluding energy under- and over-reporters before statistical analyses. Third, the score selected to measure meal quality may have some limitations for breakfast. A healthy breakfast may imply a low intake of iron-rich foods, which may decrease the overall score even though the breakfast may still meet requirements for a healthy meal (the main source of iron in breakfast in Spain would be cured meats, which were discouraged by the PREDIMED-Plus interventions for everybody). Finally, our findings only apply to older adults with excess body weight and metabolic syndrome and cannot be generalized to other populations. Despite these limitations, our research offers a novel approach to the study of the health implications of breakfast that goes beyond the mere consideration of its intake.

In conclusion, individuals at high cardiovascular risk may benefit from a balanced breakfast to maintain a healthy body weight, waist circumference, lipid profile, and renal function. A breakfast containing 20-30% of total caloric intake was linked to lower values of BMI, WC, triglycerides, and higher HDL-C concentrations, and a high-quality breakfast was associated with healthier values of WC, HDL-C, and eGFR. Our findings highlight the importance of not just eating breakfast, but paying attention to the quantity and quality of what is consumed. More studies are needed to clarify the role of breakfast quantity and quality in cardiovascular outcomes and other chronic diseases, which could help refine dietary recommendations.

## ACKNOWLEDGEMENTS

A full list of names of all PREDIMED-Plus study collaborators is available in the **Appendix**. The authors also thank the PREDIMED-Plus Biobank Network as a part of the National Biobank Platform of the ISCIII for storing and managing the PREDIMED-Plus biological samples. CIBER Fisiopatología de la Obesidad y Nutrición (CIBEROBN), CIBER Epidemiología y Salud Pública (CIBERESP), and CIBER Enfermedades Cardiovasculares (CIBERCV) are initiatives of Instituto de Salud Carlos III (Madrid, Spain), and are financed by the European Regional Development Fund.

## AUTHORS’ CONTRIBUTION

K.A.P.V.: data extraction, formal analysis, methodology, data interpretation, writing – original draft. C.L.: methodology, data interpretation, writing – review and editing. M.D.Z.: methodology, data interpretation, writing–review and editing, O.C: data interpretation, writing–review and editing. J.S.S.: funding acquisition, project administration, data interpretation, writing – review and editing. F.J.B.G.: data interpretation, writing – review and editing. D.C: funding acquisition, project administration, data interpretation, writing - review and editing. R.E.: funding acquisition, project administration, data interpretation, writing - review and editing. E.R.: funding acquisition, project administration, data interpretation, writing – review and editing. F.J.T.: funding acquisition, project administration, data interpretation, writing – review and editing. G.B.: methodology, writing – review and editing. M.M.: data interpretation, writing – review and editing. D.M.A.: methodology, writing – review and editing. H.S.: data interpretation, writing – review and editing. M.F.: study design and conceptualization, methodology, funding acquisition, project administration, data interpretation, writing – review and editing. A.H.: study design and conceptualization, data curation, formal analysis, methodology, visualization, data interpretation, writing – review and editing.

## DATA AVAILABILITY STATEMENT

The generation and analysis of the data sets within this study are not projected to be open to access beyond the core research group. This is because the participants’ consent forms and ethical approval did not include provisions for public accessibility. However, we follow a controlled data-sharing collaboration model, as the informed consent documents signed by the participants allowed for regulated collaboration with other researchers for study-related research. Following an application and approval process by the PREDIMED-Plus Steering Committee, the data described in the manuscript, alongside the codebook and analytic code, will be available upon request. Researchers interested in this study can reach out to the Committee by sending a request letter to. For those proposals that gain approval, a data-sharing agreement, outlining the specifics of the collaboration and data management, will be prepared and finalized.

